# Wearable-Measured Heart Rate Variability and Premenstrual Disorder Symptoms across Menstrual Cycle

**DOI:** 10.1101/2024.10.27.24316196

**Authors:** Qing Pan, Jing Zhou, Min Chen, Peijie Zhang, Xinyi Shi, Yifei Lin, Jin Huang, Yuchen Li, Donghao Lu

## Abstract

**IMPORTANCE:** Premenstrual disorders (PMDs), characterized by affective symptoms before menses, significantly impact women who are suffering. Current diagnostic tools are time-consuming and challenging in practice, resulting in delay in detection and treatment. There is an urgent need to identify objective and easily accessible measures to streamline the diagnostic process for PMDs.

**Objectives:** To investigate the menstrual fluctuation of wearable device-based real-time heart rate variability (HRV) through menstrual cycles and its associations with premenstrual disorders (PMD) symptoms.

**Design, Setting, and Participants:** We conducted a prospective study of female participants nested from the Care of Premenstrual Emotion Cohort.

**Exposure outcome and measures:** Daily HRV metrics (SDNN, rMSSD, LF, HF, and LF/HF) were estimated from consecutive RR-intervals (RRI) collected by the Huawei Fitness Tracker 6 Pro at 5-minute intervals over 1-2 menstrual cycles and averaged on records during 03:00-05:00 a.m. PMD symptoms were assessed with the Daily Record of Severity of Problems on a daily basis. HRV variability across cycles was described using descriptive statistics and splines, while associations between HRV metrics and PMD symptoms were estimated using a mixed-effect model.

**Results:** In total, 193 participants (with 68 prospectively confirmed PMDs) were included, with measures from 293 menstrual cycles. In both women with and without PMDs, SDNN, rMSSD, and HF decreased before menses and increased afterwards; the increase trends were more pronounced in women without PMDs. During the week before or after menses, levels of these HRV metrics were inversely associated with PMD symptoms among women with PMDs (e.g., rMSSD, postmenstrual week, β = -0.036 per SD, 95% CI: -0.048 to -0.065), whereas null association was noted for those without PMDs (β = -0.001, 95% CI -0.011 to 0.009; *P*-for-difference < 0.001). The association was particularly stronger with affective symptoms than with physiological symptoms, and more pronounced during the premenstrual week among women with premenstrual dysphoric disorder compared with those with premenstrual syndrome.

**Conclusion and Relevance:** Our findings suggest that wearable device-estimated HRV metrics fluctuate across menstrual cycles, with varying strengths of association with PMD symptoms between individuals with and without PMDs, which may aid future diagnostic process for PMDs.

**Key Points:** *Question:* Does heart rate variability (HRV) vary across menstrual cycle? Is HRV associated with premenstrual disorder symptoms in a different way between women with and without PMDs?

*Findings:* In a sample of 68 women with PMDs and 125 women without PMDs, temporal patterns across menstrual cycle were found for several wearable-measured HRV metrics in both groups. The associations between HRV and PMD symptoms during one week before or after menses were stronger among women with PMDs compared to those without.

*Meaning:* HRV fluctuates across menstrual cycles, with varying strengths of association with PMD symptoms between women with and without PMDs.

## INTRODUCTION

Premenstrual syndrome (PMS) and premenstrual dysphoric disorder (PMDD), together coined as premenstrual disorders (PMDs), affect about 20 to 30% and 2 to 8% women of reproductive age, respectively ^1^. These patients typically suffer from various affective and physical symptoms before menses, entailing functional impairments on social, relationship, and work performance ^2,3^. Our recent studies have illustrated a range of health consequences following PMDs, such as peripartum depression^4^, menopause symptoms^5^, suicidal behavior^6^, and premature death for those diagnosed in young ages ^7^. It may also contribute to the well-documented sex-disparity in mental health^8^.

Despite the high prevalence and tremendous impact, clinical challenges prevent these patients from early detection and effective treatment. On average, it takes the patients six visits to different healthcare professionals and a staggering 12 years to receive a diagnosis for PMDD ^9^. In addition to the limited awareness of PMDs among healthcare providers, the sole diagnostic tool involves maintaining a daily symptom chart for two menstrual cycles before receiving treatment. It is exceedingly time-consuming and challenging for both patients (who must diligently complete questions over two months) and clinicians (who must review extensive longitudinal data). To streamline and expedite the diagnostic process for PMDs, there is an urgent need for a diagnostic tool that utilizes objective and easily accessible measures.

Heart rate variability (HRV) has been established as a reliable non-invasive indicator of autonomic nerves system (ANS) activity ^10,11^, which has been linked several core symptoms of PMDs, e.g., anxiety, depression, and irritability ^12–14^. Given the known hormone-modulating effect on ANS activity, decreased HRV has been observed during late luteal phase among healthy women^15–18^. While these findings were based on single-timepoint or short-period measurement of HRV, the dynamic trend of HRV across the full menstrual cycle remains unclear. Moreover, few studies base on small sample size (14-29 cases ^19–22^) and varying, yet up to 24-h, recording length have found differences in HRV between women with and without PMDs ^19^ .While these findings are proactive, longitudinal measurement of HRV is needed to evaluate its potential for monitoring and diagnosing PMDs. Consumer wearables are widely used in young people and have emerged as a convenient and reliable tool for collecting real-time HRV ^23,24^. Leveraging real-time HRV from a wearable device at a minute-scale, we aimed to characterize the temporal patterns of HRV across the menstrual cycle, and assess the association of HRV with PMD symptoms between individuals with and without PMDs.

## METHODS

### Participants

The Care of Premenstrual Emotion (COPE) study is a prospective cohort of medical students at Sichuan University initiated in 2021. It aimed to collect information on demographics, health behaviors, bio-samples, and wearable device-based biomarkers of PMDs^25^. A total of 1931 female students aged 18-32 from West China Schools of Medicine (including Clinical Medicine, Stomatology, Basic Medical Sciences & Forensic Medicine, Public Health, and Pharmacy) were recruited. The COPE study was approved by the Institutional Review Board of West China Hospital, Sichuan University (No. 2018-535). All participants were fully informed about the purpose and content of the study and provided their electronic consent to participate.

The present study was based on a sub-population of natural cyclers from the COPE cohort. Participants were initially screened for PMDs using a modified version of the Calendar of Premenstrual Symptoms scale. This scale includes 8 affective and 19 physical/behavioral symptom items, followed by 3 functional impact items, all measured on a 4-point Likert scale. The classification criteria have been described in our previous study^25^. All participants who screened positive for PMDs were invited, while a random sample of those who screened negative were recruited (Supplementary Methods). They were asked to wear the tracker for at least one cycle, and complete prospective symptom charting for two cycles (one cycle for those screened negative). Individuals with severe somatic diseases or neurological disorders were not invited. After excluding 9 withdraws and 2 with a high missing rate on prospective symptom charting, a total of 193 participants were included for analysis.

### HRV measurements

All participants were instructed to wear a Huawei Fitness Tracker 6 Pro for at least 12 hours each day throughout their menstrual cycles and were required to upload their daily records to a cloud-based service. The fitness tracker is a wristband equipped with photoplethysmography (PPG) sensor, it automatically collect real-time metrics, including RRI, heart rate, skin temperature, blood oxygen saturation, physical activity, and sleep episodes. HRV metrics measured the variability of RRI, which were proved to be comparable to these abstained from medical electrocardiogram (ECG) ^24^. After removing outliers with heart rate beyond 20-200 beats/min, two time-domain parameters, including SDNN (Standard Deviation of NN intervals) and rMSSD (Root Mean Square of Successive Differences between NN intervals) were calculated. Then non-uniformly distributed RRI records were further transformed into uniformly distributed records at 4Hz (four records per minute). This transformation employs linear interpolation to estimate RRI values at uniformly distributed time points. Then three frequency-domain parameters, including LF (absolute power in the low-frequency band, 0.04–0.15 Hz), HF (absolute power in the high-frequency band, 0.15–0.4 Hz), and LF/HF (the ratio of LF to HF) were calculated by Fourier transformation.

### Assessment of PMD symptoms

Participants were asked to complete a questionnaire daily concerning PMD symptoms and record menstruation occurrence. PMD symptoms were assessed with a Chinese version^26^ of Daily Record of Severity of Problems (DRSP)^27^, which has been proven to be both sensitive and reliable in measuring symptoms and impairment for PMDs^27^. The DRSP consists of 24 items, each rated on a 6-point Likert scale ranging from 1 (not at all) to 6 (extreme). For each individual item, the average values of before (day -5 to -1) and after (day 6 to 10) menses were calculated to estimate the symptom change from pre- to post-menstrual period. In line with established criteria^28,29^, individuals were diagnosed with PMDs if they met the following criteria: 1) at least one core symptom scores >2 on average in premenstrual days (Day -5 to -1) and with 20% decrease in postmenstrual days (Day 6 to 10); 2) in addition, at least four symptom scores >1 in premenstrual days and with 20% decrease in postmenstrual days. 3) at least one functional impairment scores >2 in premenstrual days. Among the PMD cases, individuals were further classified as PMS and PMDD; the latter was defined as 1) at least one core symptom scores >3 on average in premenstrual days and with 20% decrease in postmenstrual days; 2) at least four symptom scores >2 in premenstrual days and with 20% decrease in postmenstrual days. 3) a least one functional impairment scores >3 in premenstrual days. The definitions of core symptoms were listed in Supplement Methods.

### Covariates

Participants provided information on their age and schools at recruitment, and risk factors of PMDs, e.g., age of menarche, body mass index (BMI; calculated from self-reported height and weight), and alcohol consumption (having consumed alcohol at least once, or heavy drinking, defined as consuming five or more drinks within one to two hours, during the past 30 days).

Smoking status was not included as a covariate due to its low prevalence (1.85%) among young women in China^30^.

### Statistical analysis

We estimated the average levels of the five HRV metrics (including mean and standard deviation for normally distributed parameters and the median and percentiles for non-normally distributed parameters) by days across menstrual cycle among individuals with and without PMDs. To visualize the daily fluctuations, we applied natural cubic splines with two knots at day -5 and day 8, spaced at 33% and 67% quantiles of menstrual days. Moreover, we employed a mixed effect model to estimate the weekly association between HRV and PMD symptoms, because both HRV metrics and PMD symptoms were repeatedly measured. The weeks were defined as two weeks before (days -14 to -8), one week before (days -7 to -1), one week after (days 1 to 7), and two weeks after menses (days 7 to 14). *Z* score test was used to compare the association strength between individuals with and without PMDs across menstrual weeks.

To illustrate the relationship with different PMD symptom domains, we assessed the associations of HRV metrics with physiological/behavioral and affective symptom scores (z-score), separately. Definitions of the physiological/behavioral and affective symptom scores can be found in the Supplement Methods. To shed light on the disease severity, we also assessed the association between HRV and PMD symptoms between PMDD versus PMS. Since our samples included sub-clinical PMDs (assessed as controls), we conducted a sensitivity analysis restricting the controls to “pure controls” (i.e., those with minimum PMD symptoms). The pure controls were defined as reporting ‘not at all’ or ‘minimal’ for all individual symptoms and the impact questions. Last, sensitivity analysis was performed in subpopulations reporting regular menstrual cycles with a length of 26 to 31 days.

All analyses were conducted using R statistical software (Version 4.3.2; R Core Team, 2023), with significance set at α = 0.05. A *P*-value less than or equal to 0.05 was considered statistically significant.

## RESULTS

### Characteristics

A total of 193 participants with daily records across 293 menstrual cycles were included in analysis, with an average age of 19.92±1.65 years. Among them, 68 (35.2%) were prospectively diagnosed with PMDs, including 41 (21.2%) with PMS and 27 (14.0%) with PMDD. Individuals with PMDs were statistically comparable to those without regarding age, age at menarche, BMI, and alcohol consumption (all *P* > 0.05; Table 1).

**Table 1.** Characteristics between individuals with PMDs and without PMDs.

| **Characteristics** | **Without PMDs**<br>**(N = 125)** | **PMDs**<br>**(N = 68)** | **P value** |
| --- | --- | --- | --- |
| **Age (years)** | 19.82 ± 1.41 | 20.09 ± 2.00 | 0.286 |
| **Age at menarche (years)** | 12.34 ± 1.22 | 12.31 ± 1.25 | 0.866 |
| **BMI (kg/m<sup>2</sup>)** | 21.14 ± 4.59 | 20.79 ± 2.43 | 0.564 |
| **Drinking during the past 30 days** |  |  | 0.246 |
| 0 day | 57 (46) | 23 (34) |  |
| 1-2days | 26 (21) | 19 (28) |  |
| >3 days | 41 (33) | 26 (38) |  |
| **Drinking heavily during the past 30 days** |  |  | 0.469 |
| 0 day | 76 (61) | 44 (65) |  |
| 1-2days | 14 (11) | 4 (6) |  |
| >3 days | 34 (27) | 20 (29) |  |

Note: PMDs: premenstrual disorders, including PMS and PMDD. BMI: Body mass index; Drinking heavily: consuming at least 5 drinks within one or two hours.

### PMD symptom and HRV levels across menstrual cycle

Among individuals with PMDs, a bell-shape pattern was observed for PMD symptom across menstrual cycle, with the peak at the day before menses (Day -1; Figure 1A). In contrast, a relatively flat pattern with lower symptom score was found for those without PMDs. Regarding HRVs, we observed a decrease of SDNN in early luteal phase (from Day -13 to -6) and an increase from Day -5 until Day 11 (Figure 1B) for both individuals with and without PMDs, with a more pronounced elevation among those without PMDs. Similar patterns were found for rMSSD and HF (Figure 1C/E). The variation pattern for LF and LF/HF ratio was largely similar between those with and without PMDs (Figure 1D/F) .

**Figure 1.**
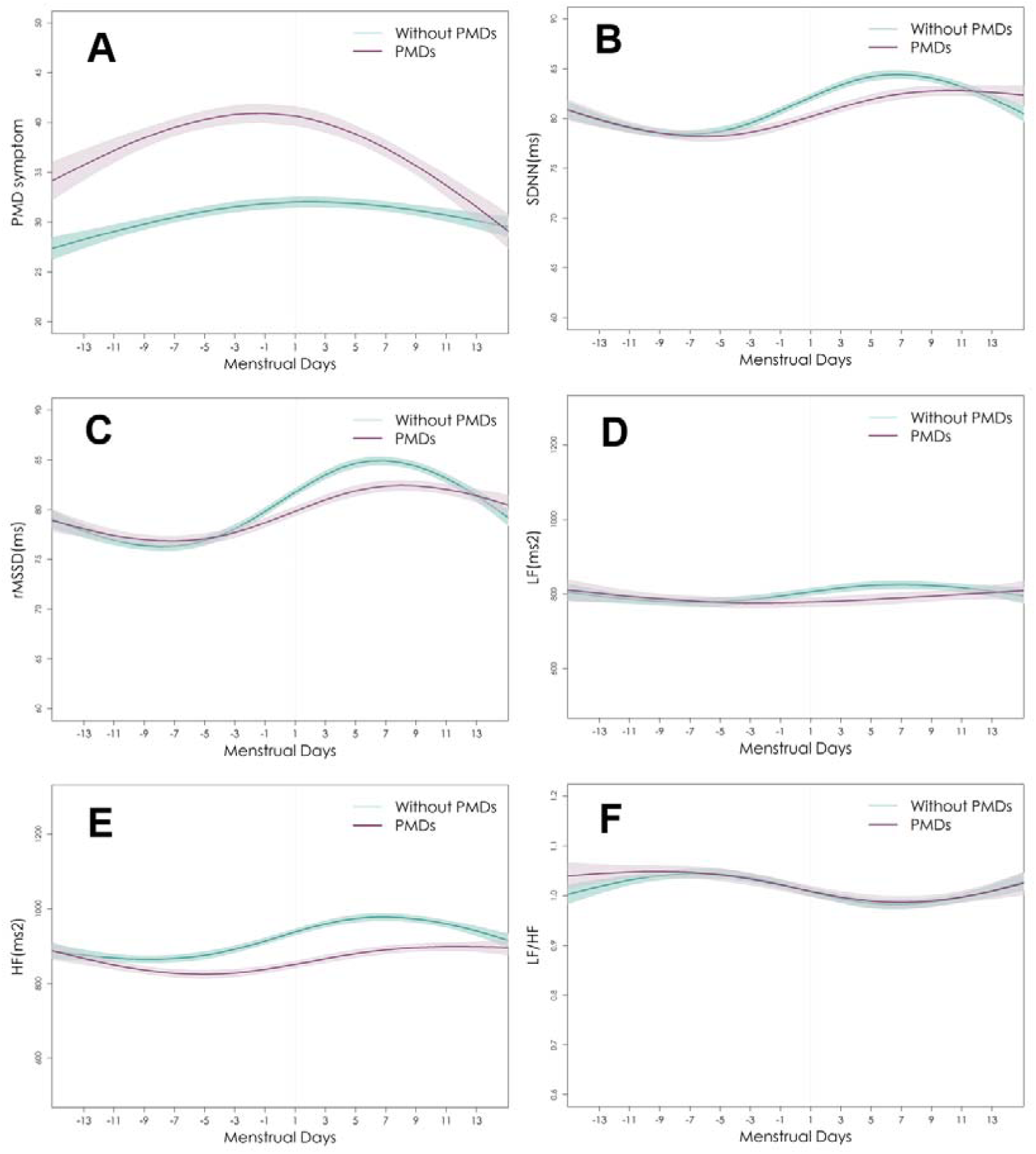
Comparisons of PMDs symptom score and HRV levels across menstrual cycle between individuals with and without PMDs (Note: PMDs: premenstrual disorders, including PMS and PMDD. A: PMD symptom: Symptom scores for premenstrual disorders were assessed using the Daily Record of Severity of Problems scale; B: SDNN: standard deviation of RR intervals; C: rMSSD: Root mean square of successive RR interval differences; D: LF: absolute power in the LF band (0.04–0.15 Hz); E: HF: absolute power in the HF band (0.15–0.4 Hz); F: LF/HF: ratio of LF to HF.)

### Associations between HRVs and PMD symptom

While we presented the associations over four weeks across menstrual cycle, we focused on results during the week before (symptom window) and the week after menses (symptom-relieving window). During the week before menses, the SDNN level was inversely associated with PMD symptom among individuals with PMDs (β = -0.024 per SD increase, 95% CI -0.036- -0.041; Figure 2A and point estimates in Table S1), whereas null association was noted among those without PMDs (β = 0.001, 95% CI -0.007-0.009; *P*-for-difference < 0.001).

**Figure 2.**
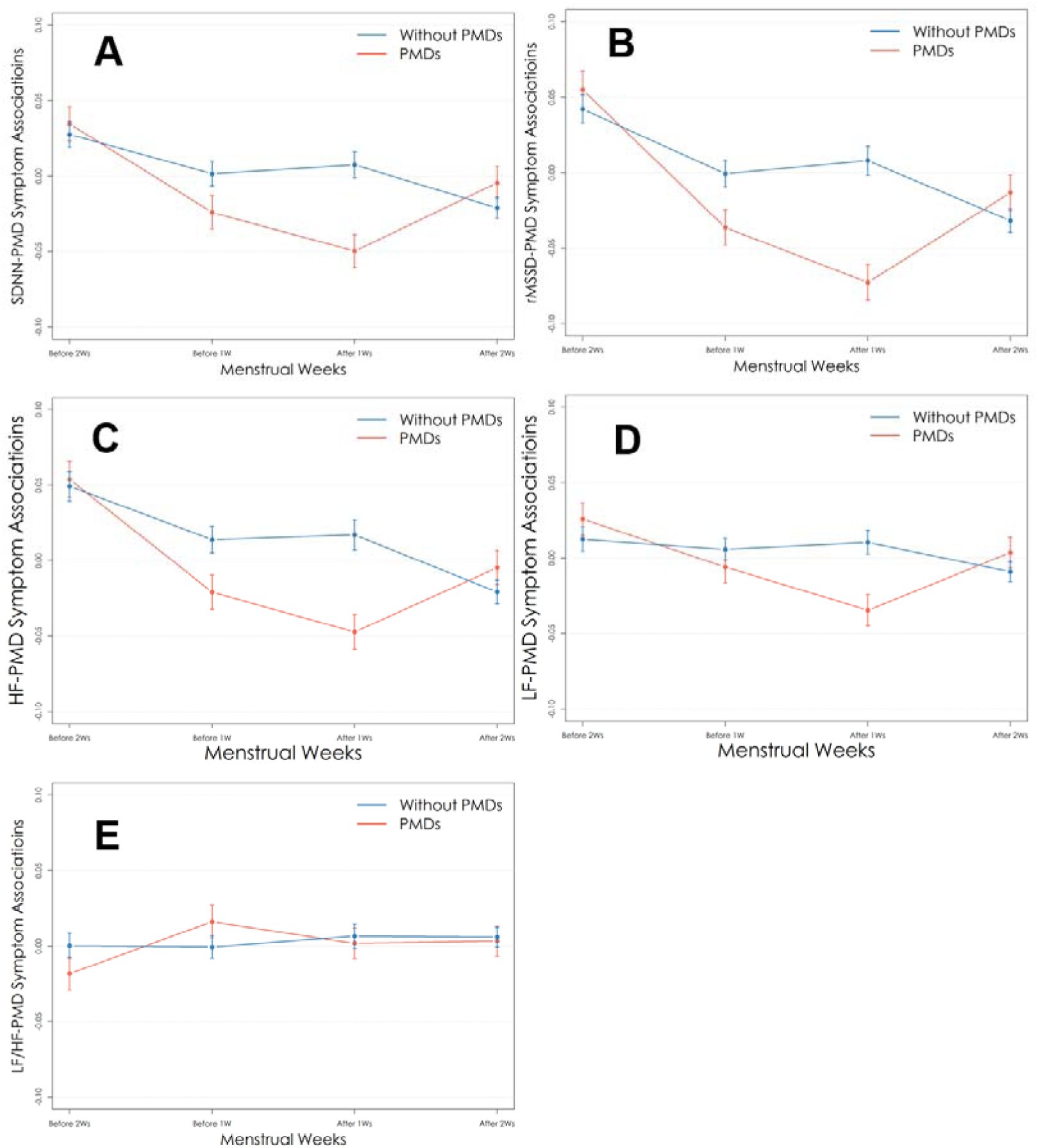
Comparisons of the associations between HRVs and PMD symptoms among individuals with PMDs and without PMDs across menstrual cycle (Note: PMDs: premenstrual disorders, including PMS and PMDD. A: SDNN: standard deviation of RR intervals; B: rMSSD: Root mean square of successive RR interval differences; C: HF: absolute power in the HF band (0.15–0.4 Hz); D: LF: absolute power in the LF band (0.04–0.15 Hz); E: LF/HF: ratio of LF to HF. The models have been adjusted for age, age at menarche, BMI, drinking during the past 30 days, and drinking heavily during the past 30 days.).

Even stronger association difference between those with and without PMDs was found during the week after menses (*P*-for-difference < 0.001). Largely comparable trends were also noted for rMSSD, HF, and LF (Figure 2B, 2C, 2D) but not observed for LF/HF (Figure 2E).

### Additional analyses

When analyzing physiological/behavioral and affective symptoms separately, we observed similar patterns found in the primary analysis (Figure 3 and estimates in Table S2), yet stronger difference for affective symptoms between individuals with and without PMDs.

**Figure 3.**
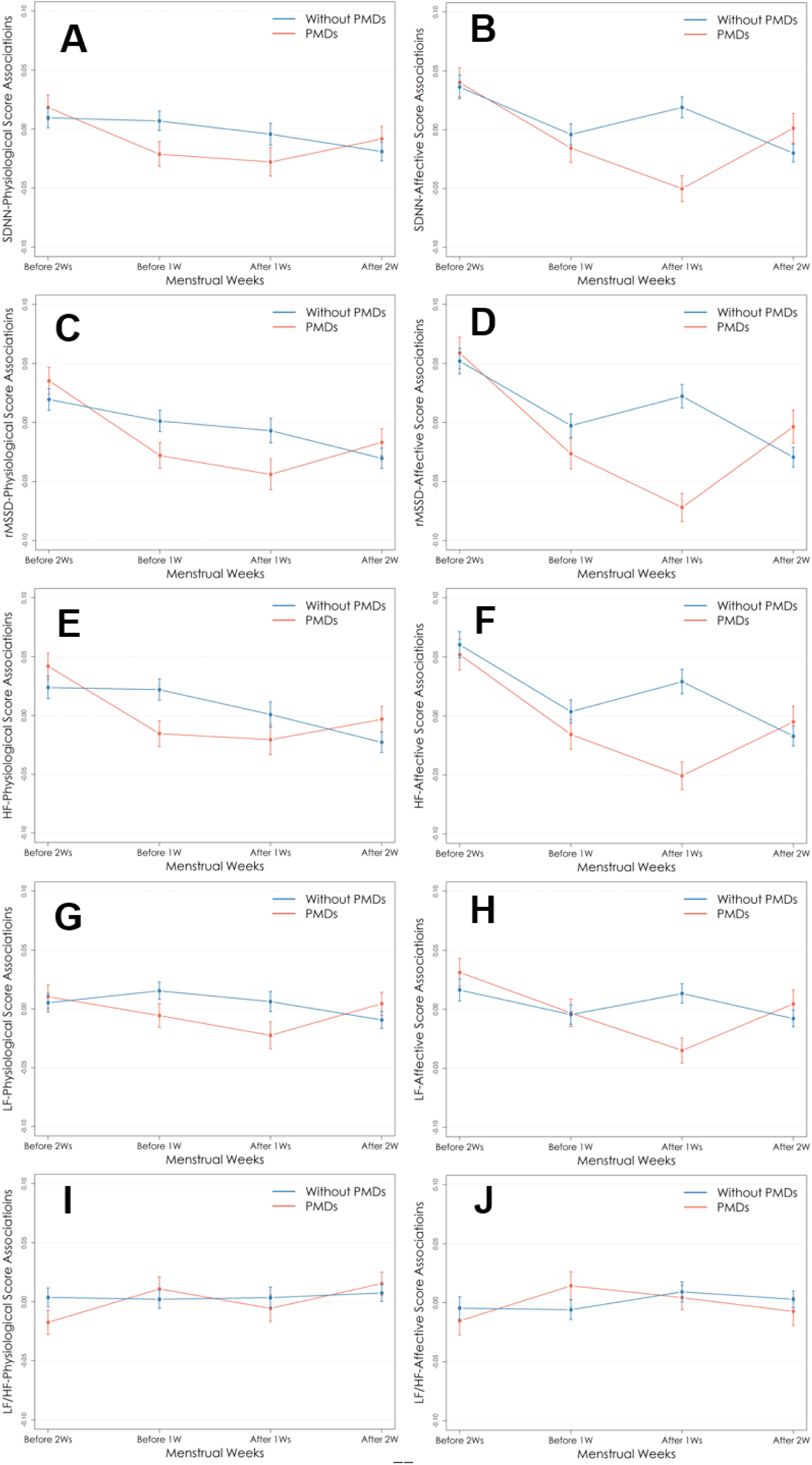
Comparisons of the associations between HRV and physiological/behavioral and affective symptoms among individuals with PMDs and without PMDs across menstrual cycle. (Note: PMDs: premenstrual disorders, including PMS and PMDD. PMDD: premenstrual dysphoric disorder; PMS: premenstrual syndrome; A: SDNN for physiological/behavioral sub-scores; B: SDNN for affective sub-scores; C: rMSSD for physiological/behavioral sub-scores; D: rMSSD for affective sub-scores; E: HF for physiological/behavioral sub-scores; F: HF for affective sub-scores; G: LF for physiological/behavioral sub-scores; H: LF for affective sub-scores; I: LF/HF for physiological/behavioral sub-scores; J: LF/HF for affective sub-scores. The models have been adjusted for age, age at menarche, BMI, drinking during the past 30 days, and drinking heavily during the past 30 days.)

Regarding PMD severity, we found that the inverse association of SDNN were comparable between PMS and PMDD groups during the week after menses (*P*-for-difference 0.868), while a stronger association was found for PMDD within the week before menses (*P*-for-difference 0.006). Similar heterogeneous associations were also observed for rMSSD, HF, and LF (Figure 4 and Table S3). In addition, we compared PMDs with a pure control group consisting of 43 individuals and yielded comparable trends (Figure S1 and Table S4). Last, we restricted the analysis to individuals with regular menstrual cycles with a cycle length of 26 to 31 days and observed similar results (Figure S2-3 and Table S5).

**Figure 4.**
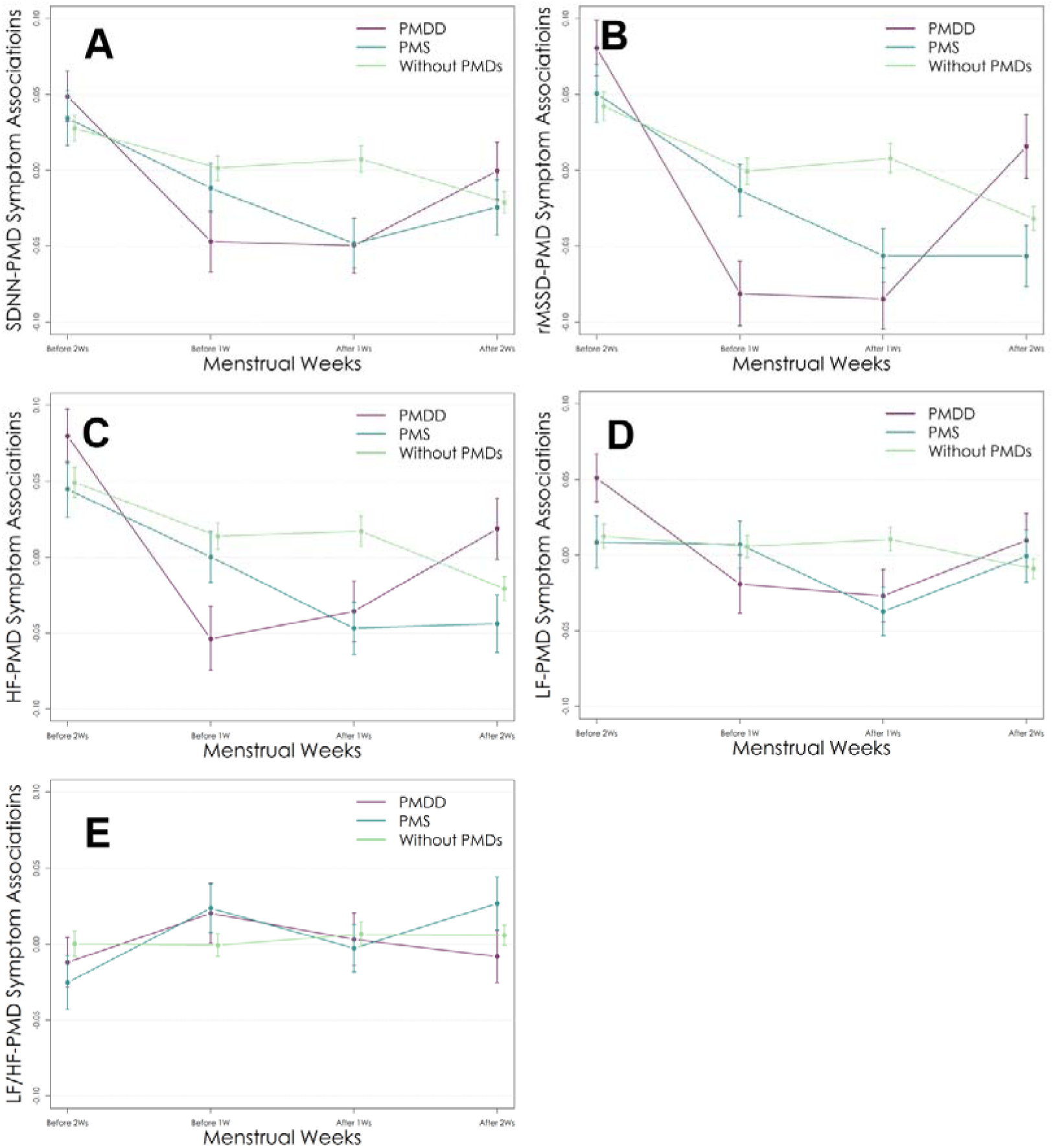
Comparison of the associations between HRV and PMD symptoms among individuals with PMDD, PMS, and without PMDs across the menstrual cycle (Note: PMDs: premenstrual disorders, including PMS and PMDD; PMS: premenstrual syndrome; PMDD: premenstrual dysphoric disorder. Non-PMDs: individuals without PMDD or PMS. A: SDNN: standard deviation of RR intervals; B: rMSSD: Root mean square of successive RR interval differences; C: HF: absolute power in the HF band (0.15–0.4 Hz); D: LF: absolute power in the LF band (0.04–0.15 Hz); E: LF/HF: ratio of LF to HF. The models have been adjusted for age, age at menarche, BMI, drinking during the past 30 days, and drinking heavily during the past 30 days.)

## Discussion

To the best of our knowledge, this study is the first investigation into the real-time dynamics of HRV across menstrual cycles and its associations with PMD symptoms. We showed that measuring with wearable device, most HRV indicators fluctuated across the menstrual cycle in both healthy women and women with PMDs, with more pronounced alteration from weeks before menses to weeks after menses among healthy women. Our findings highlight that information on menstrual cycle phase should be considered in future HRV studies involving women. Moreover, HRV was inversely associated with PMD symptoms during the week before and after menses among women with PMDs, whereas no such link was found for healthy women. The association was particularly stronger with affective symptoms and during the premenstrual week among women with PMDD. Given the common use of wearable device and easy access to passively collected HRV readouts, our findings suggest that HRVs can serve as valuable predictive markers aiding risk stratification and diagnosis of PMDs.

### HRV across menstrual cycles

Previous studies based on single-time or short-period measurement have documented that HRV indicating parasympathetic activity (SDNN, rMSSD, and HF) decreases during luteal phase compared to follicular phase in healthy women ^15–18,31^. With a real-time monitoring through wearable devices, our study not only corroborated the previous findings but also captured the dynamic changes of HRVs across menstrual cycle. Specifically, parasympathetic activity indicated by SDNN, rMSSD, and HF decreased during early luteal phase, with the lowest level about one week before menses before it bounced back and peaked around one week after menses. Additionally, we observed that the LF/HF ratio (representing the balance between sympathetic and parasympathetic activities, where a lower value indicates parasympathetic dominance and a higher value suggests sympathetic dominance ^32^) showed a slight decrease before menses, followed by an increase afterward. This may be explained by the fact that during the premenstrual phase, as parasympathetic activity decreases, sympathetic activity gradually becomes dominant. After menstruation, parasympathetic activity regains dominance. Hormonal shifts during menstrual cycle might explain these HRV fluctuations. The rise in estrogen levels after menses enhances parasympathetic activity and suppresses sympathetic outflow, thereby contributing to elevated SDNN, rMSSD, and HF values^33^. Conversely, the peak in progesterone levels in luteal phase attenuates baroreflex sensitivity and augments sympathetic activation ^21,34^, which is reflected in increased LF/HF values.

Limited evidence is available on HRV trajectory across menstrual cycle among individuals with PMDs. In a US study of 9 cases and 12 controls with selected two specific days from follicular and luteal phases respectively^20^, lower SDNN, rMSSD, and HF values were noted during the late luteal phase among women with PMDs compared to those without, which is in line with our finding. However, we found that during most post-menstruation days (corresponding to the follicular phase in the previous study), SDNN, rMSSD, and HF remained lower in individuals with PMDs when compared with healthy controls, whereas the previous study reported these values as being higher in the PMS group compared to controls. While the discrepancy might be explained by the small sample size and limited measurement length in the previous study, future studies are warranted to confirm our findings. If replicated, our results suggest that the ability of restoring parasympathetic activity from late luteal to early follicular phase is impaired among women with PMDs.

### HRV and PMD symptoms

To our knowledge, there are only two studies investigating the association between HRV and PMD symptoms. In a Polish study with 177 women of reproductive age, including both patients with PMDs and healthy controls, Danel et al. found a positive relationship between PMD symptoms and HRV indices (SDNN and rMSSD, measured within 10 minutes intervals) during follicular phase in women without use of hormonal contraception, but not for those with use of hormonal contraception^35^. Another study tracking three participants throughout the entire menstrual cycle reported that, within individuals, log(rMSSD) (measured within 6 minutes intervals) was inversely correlated with psychological symptom but showed no association with physiological symptom ^36^. While the majority of our participants did not use hormonal contraceptives (95.5%), our study significantly differs from previous ones by design (larger sample size and consecutive measurements on both HRV and PMD symptoms), and unsurprisingly yielded different findings. For instance, we observed that in one week before or after menses, rMSSD levels were inversely associated with PMD symptoms among individuals with PMDs, whereas no significant association was found for those without PMDs. The association was also observed for SNDD and HF, and it was particularly stronger during the one week after menses. Our findings on the heterogeneous association between HRV and PMD symptoms around menses between women with and without PMDs suggest that HRV metrics may serve as potential biomarkers to aid the diagnostic process for PMDs.

Although the exact mechanism linking ANS activity to PMDs remains unclear^37–40^, the prevailing theory supports a role of hormonal fluctuations^41,42^. During luteal phase, fluctuations in estrogen and progesterone can disrupt ANS balance by affecting neurotransmitters such as norepinephrine and serotonin^43^. Increased sympathetic nervous activity has been associated with anxiety, irritability, and other emotional symptoms, while reduced parasympathetic activity is linked to difficulties in emotional regulation^44^. On the other hand, ANS regulates physiological functions such as blood pressure, heart rate, and HRV. Specifically, certain HRV values would increase as parasympathetic effect increases, while HRV would decrease as sympathetic effect increases ^45^. The relationship between HRV and PMD symptoms observed in our data may therefore speak for the phenotypic link underlying ANS activity. This is further supported by our data showing that the association between HRV and affective symptoms was stronger than for physiological/behavioral symptoms.

### Strengths and limitations

The strength of this study lies in real-time monitoring of HRV, coupled with prospective PMD symptom charting in a relatively large sample, providing a panoramic picture of autonomic function and PMD symptoms across menstrual cycles. However, several limitations need consideration. First, the PPG sensor in the wristband may be affected by motion artifacts and variations in fit tightness, potentially leading to inaccuracy in HRV readings. However, it has been validated that HRV metrics collected by PPG sensor from Huawei fitness tracker are comparable to those obtained from medical ECGs ^24^.Moreover, we calculated HRV during the resting state by averaging records collected between 3:00 and 5:00 a.m., when participants are less likely to move. Second, although PMD symptoms were assessed daily through validated electronic survey and PMD cases were confirmed based on established criteria, self-reported symptoms may introduce inaccuracy due to variations in individual interpretation. However, the temporal pattern of PMD symptoms were assessed within the same individual for diagnosing PMDs. In addition, our study aimed to assess the predictive value of HRV for PMD symptoms instead of claiming causal relationship, although a few known confounders were adjusted for in our analyses. Last, the study’s sample was based on health professional students, which may limit the generalizability of the findings to broader populations.

## Conclusion

In summary, our findings suggest that HRV metrics fluctuate across the menstrual cycle in healthy women, highlighting that menstrual cycle phase should be considered in future HRV studies involving women. Our findings also illustrate that several HRV metrics are only inversely associated with PMD symptoms during the week before and after menses among women with PMDs. While future studies based on independent populations are needed to confirm the predictive value, these proactive findings illuminate that HRV metrics could play a crucial role in improving the diagnostic process for PMDs in the future.

## Data Availability

All data produced in the present study are available upon reasonable request to the authors

## Acknowledgement

This work was supported by the National Natural Science Foundation of China (No. 81801359 and 82271581 to Dr. Lu) and National Natural Science Foundation of Sichuan Province (No. 2023NSFSC1487 to Dr. Li). Dr. Lu has also received support from the Swedish Research Council for Health, Working Life and Welfare (FORTE) (No. 2020-00971 and 2023-00399), Swedish Research Council (2020-01003), and Karolinska Institutet. Most importantly, we thank all the participants in the COPE cohort for their support and patience.

## Author Contributions

Qing Pan and Donghao Lu had full access to all of the data in the study and takes responsibility for the integrity of the data and the accuracy of the data analysis.

*Concept and design:* Donghao Lu.

*Acquisition, analysis, or interpretation of data:* Qing Pan, and Jing Zhou

*Drafting of the manuscript:* Qing Pan. *Critical revision of the manuscript for important intellectual content:* Donghao Lu, Yuchen Li, and Jing Zhou. *Statistical analysis:* Qing Pan. *Obtained funding:* Donghao Lu. *Administrative, technical, or material support:* Min Chen, Peijie Zhang, and Xinyi Shi. *Supervision:* Donghao Lu.

## Data Sharing Statement

The data presented in this study is available from the corresponding author on reasonable request.

## Additional Contributions

We are grateful to thank all the participants in the COPE cohort for their support and patience.

## Additional Information

None.

