## Supplementary for "Wearable-Measured Heart Rate Variability and Premenstrual Disorder Symptoms across Menstrual Cycle"

**Supplementary Methods**

**Sampling**

In the present study, all participants who screened positive for PMDs were invited, while those screened negative were randomly selected and underrepresented. To represent the distribution from the full cohort, participants were weighed on their selection probability in all analyses. Specifically, individuals who were screened positive were assigned a weight of 0.664 (0.234/0.352), and those screened negative were assigned a weight of 1.182 ((1 - 0.234)/(1 - 0.352)). The weights of 0.234 and 0.352 were the proportions of PMDs in the COPE cohort and current subpopulation, respectively.

**Definition of physiological/behavioral and affective symptoms**

The DRSP scale included 24 items: 1) Felt depressed, sad, down, or blue; 2) Felt hopeless; 3) Felt worthless or guilty; 4) Felt anxious, tense, keyed up, or on edge; 5) Had mood swings (e.g., suddenly felt sad or tearful); 6) Was more sensitive to rejection or feelings were more easily hurt; 7) Felt angry, irritable; 8) Had conflicts or problems with people; 9) Had less interest in usual activities (e.g., work, school, friends, hobbies); 10) Had difficulty concentrating; 11) Felt lethargic, tired, fatigued, or had a lack of energy; 12) Had increased appetite or overate; 13) Had cravings for specific foods; 14) Slept more, took naps, found it hard to get up when intended; 15) Had trouble getting to sleep or staying asleep; 16) Felt overwhelmed or that I could not cope; 17) Felt out of control; 18) Had breast tenderness; 19) Had breast swelling, felt bloated, or had weight gain; 20) Had headache; 21) Had joint or muscle pain; 22) At work, school, home, or in daily routine.at least one of the problems noted above caused reduced productivity or inefficiency; 23) At least one of the problems noted above interfered with hobbies or social activities (e.g., avoided or did less); 24) At least one of the problems noted above interfered with relationships with others. Sub-items including core symptoms, functional impairment, physiological/behavioral symptoms, and affective symptoms were defined as follow:

| No | Sub-scores | Items |
|---|---|---|
| 1 | Core symptoms | 1-8 |
| 2 | Functional impairment | 22-24 |
| 3 | Affective symptoms | 1-9, 16, and 17 |
| 4 | Physiological/behavioral symptoms | 10-15, and 18-21 |

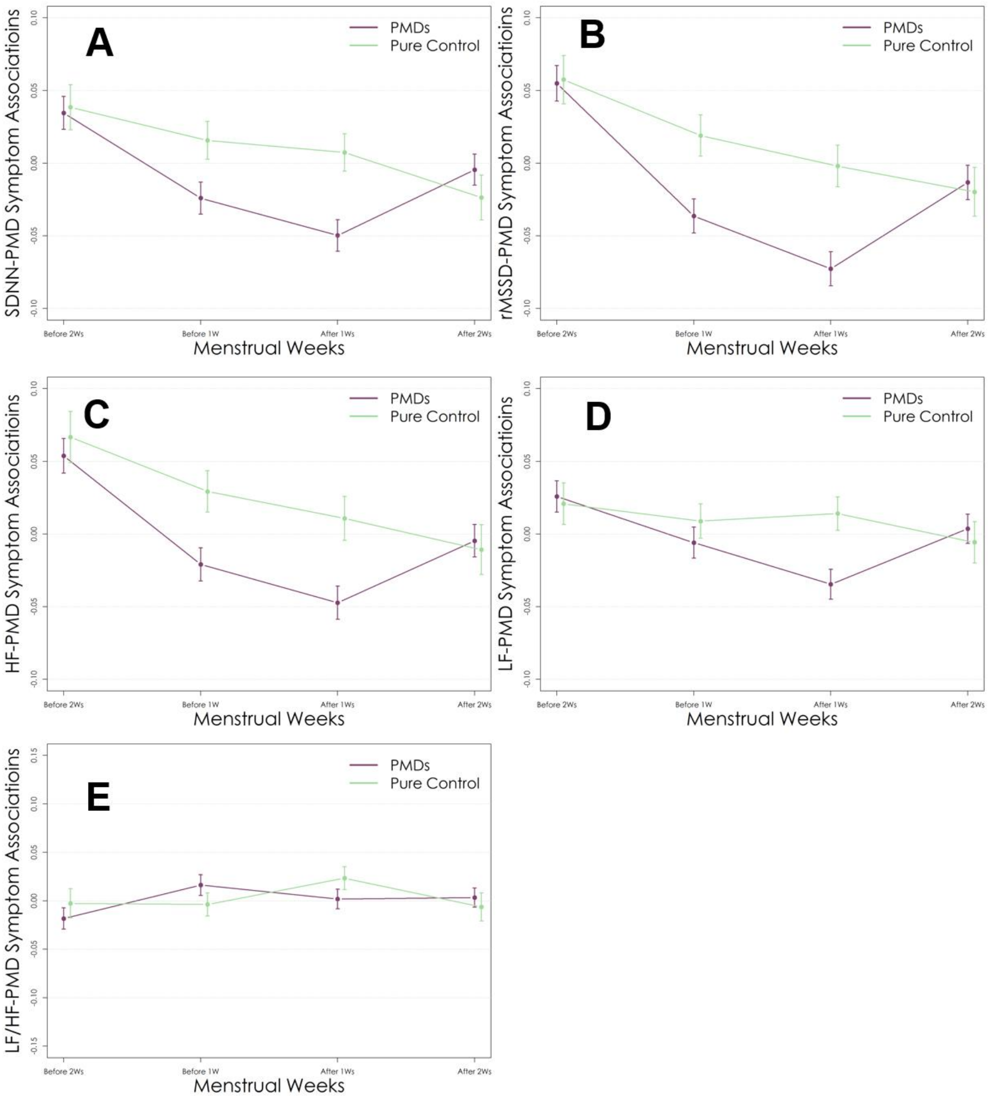


**Figure S1. Comparison of the associations between HRV and PMD symptoms among individuals with PMDs and pure control groups across the menstrual weeks** (Note: PMDs: premenstrual disorders, including PMS and PMDD. PMDD: premenstrual dysphoric disorder; PMS: premenstrual syndrome; . A: SDNN: standard deviation of RR intervals; B: rMSSD: Root mean square of successive RR interval differences; C: HF: absolute power in the HF band (0.15–0.4 Hz); D: LF: absolute power in the LF band (0.04–0.15 Hz); E: LF/HF: ratio of LF to HF. The models have been adjusted for age, age at menarche, BMI, drinking during the past 30 days, and drinking heavily during the past 30 days.)

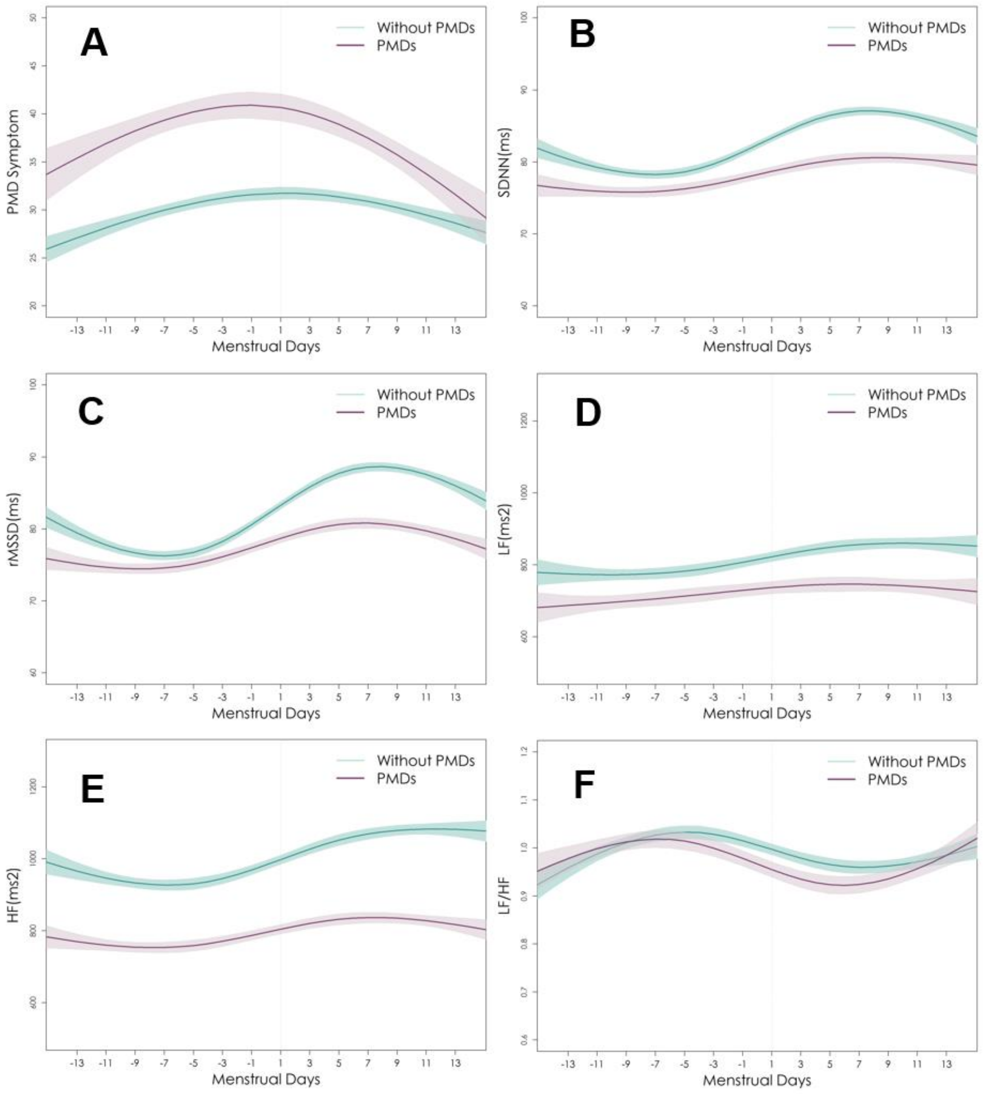


**Figure S2. Comparisons of PMD symptom score and HRV levels across menstrual days between individuals with and without PMDs in subpopulation with regular menstrual cycles** (Note: PMDs: premenstrual disorders, including PMS and PMDD. A: PMD symptom: Symptom scores for premenstrual disorders were assessed using the Daily Record of Severity of Problems scale; B: SDNN: standard deviation of RR intervals; C: rMSSD: Root mean square of successive RR interval differences; D: LF: absolute power in the LF band (0.04–0.15 Hz); E: HF: absolute power in the HF band (0.15–0.4 Hz); F: LF/HF: ratio of LF to HF. )

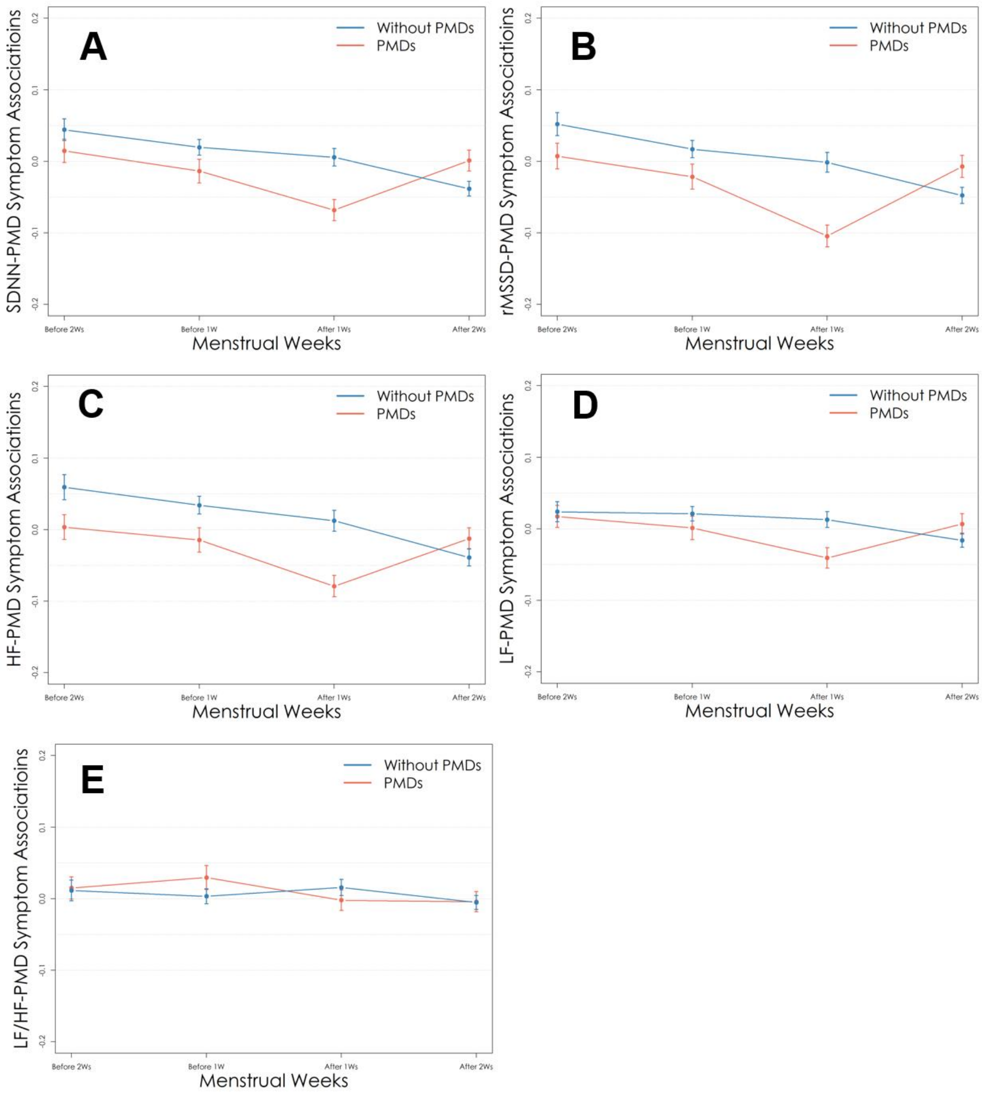


**Figure S3. Comparisons of the associations between HRVs and PMD symptom among individuals with PMDs and without PMDs across menstrual weeks in subpopulation with regular menstrual cycles** (Note: PMDs: premenstrual disorders, including PMS and PMDD. A: SDNN: standard deviation of RR intervals; B: rMSSD: Root mean square of successive RR interval differences; C: HF: absolute power in the HF band (0.15–0.4 Hz); D: LF: absolute power in the LF band (0.04–0.15 Hz); E: LF/HF: ratio of LF to HF. The models have been adjusted for age, age at menarche, BMI, drinking during the past 30 days, and drinking heavily during the past 30 days.)

**Table S1. Comparisons of the associations between HRVs and PMD symptoms among individuals with PMDs and without PMDs across menstrual cycle**

| HRVs | *With PMDs* | | *Without PMDs* | | *P-for-difference* |
|---|---|---|---|---|---|
| | ***$B_1$* ( 95%CI)** | ***P value*** | ***B2* ( 95%CI)** | ***P value*** | |
| **SNDD** | | | | | |
| Two weeks before | 0.035(0.023,0.075) | <0.001 | 0.028(0.02,0.036) | <0.001 | 0.337 |
| One week before | -0.024(-0.036,-0.041) | <0.001 | 0.001(-0.007,0.009) | 0.720 | <0.001 |
| One week after | -0.050(-0.062,-0.092) | <0.001 | 0.007(-0.001,0.015) | 0.093 | <0.001 |
| Two weeks after | -0.004(-0.014,-0.003) | 0.411 | -0.021(-0.029,-0.013) | <0.001 | 0.011 |
| **rMSSD** | | | | | |
| Two weeks before | 0.055(0.043,0.114) | <0.001 | 0.042(0.032,0.052) | <0.001 | 0.105 |
| One week before | -0.036(-0.048,-0.065) | <0.001 | -0.001(-0.011,0.009) | 0.883 | <0.001 |
| One week after | -0.073(-0.085,-0.137) | <0.001 | 0.008(-0.002,0.018) | 0.106 | <0.001 |
| Two weeks after | -0.013(-0.025,-0.019) | 0.028 | -0.032(-0.04,-0.024) | <0.001 | 0.010 |
| **HF** | | | | | |
| Two weeks before | 0.054(0.042,0.112) | <0.001 | 0.049(0.039,0.059) | <0.001 | 0.557 |
| One week before | -0.021(-0.033,-0.035) | <0.001 | 0.014(0.006,0.022) | 0.002 | <0.001 |
| One week after | -0.047(-0.059,-0.086) | <0.001 | 0.017(0.007,0.027) | <0.001 | <0.001 |
| Two weeks after | -0.005(-0.017,-0.004) | 0.414 | -0.021(-0.029,-0.013) | <0.001 | 0.021 |
| **LF** | | | | | |
| Two weeks before | 0.026(0.016,0.056) | <0.001 | 0.012(0.004,0.02) | 0.003 | 0.052 |
| One week before | -0.006(-0.016,-0.007) | 0.274 | 0.006(-0.002,0.014) | 0.127 | 0.078 |
| One week after | -0.035(-0.045,-0.064) | <0.001 | 0.010(0.002,0.018) | 0.010 | <0.001 |
| Two weeks after | 0.004(-0.006,0.013) | 0.480 | -0.009(-0.015,-0.003) | 0.007 | 0.039 |
| **LF/HF** | | | | | |
| Two weeks before | -0.018(-0.03,-0.029) | 0.001 | 0.000(-0.008,0.008) | 0.940 | 0.008 |

| | | | | | |
|---|---|---|---|---|---|
| One week before | 0.016(0.006,0.036) | 0.003 | -0.001(-0.009,0.007) | 0.868 | 0.011 |
| One week after | 0.002(-0.008,0.009) | 0.725 | 0.007(-0.001,0.015) | 0.111 | 0.469 |
| Two weeks after | 0.003(-0.007,0.011) | 0.516 | 0.006(0.000,0.012) | 0.081 | 0.676 |

(**Note:** PMDs: premenstrual disorders, including PMS and PMDD. $B_1$: the coefficient of HRV-symptom associations among these with PMDs; $SE_1$: standardized error for $B_1$; $B_2$: the coefficient of HRV-symptom associations among these without PMDs; $SE_2$: standardized error for $B_2$; *P*-for-difference: *P* value for comparisons of $B_1$ and $B_2$. SDNN: standard deviation of RR intervals; rMSSD: Root mean square of successive RR interval differences; HF: absolute power in the HF band (0.15–0.4 Hz); LF: absolute power in the LF band (0.04–0.15 Hz); LF/HF: ratio of LF to HF. The models have been adjusted for age, age at menarche, BMI, drinking during the past 30 days, and drinking heavily during the past 30 days.).

**Table S2. Comparisons of the associations between HRV and physiological/behavioral and affective symptoms among individuals with PMDs and without PMDs across menstrual cycle**

| Symptom domain | HRVs | *With PMDs* | | *Without PMDs* | | *P-for-difference* |
|---|---|---|---|---|---|---|
| | | ***$B_1$* ( 95%CI)** | ***P value*** | ***B2* ( 95%CI)** | ***P value*** | |
| Physiological/behavioral symptoms | **SNDD** | | | | | |
| | Two weeks before | 0.018(0.008,0.028) | 0.001 | 0.010(0.002,0.018) | 0.024 | 0.209 |
| | One week before | -0.021(-0.031,-0.011) | <0.001 | 0.007(-0.001,0.015) | 0.096 | <0.001 |
| | One week after | -0.028(-0.04,-0.016) | <0.001 | -0.004(-0.014,0.006) | 0.365 | 0.002 |
| | Two weeks after | -0.008(-0.018,0.002) | 0.122 | -0.019(-0.027,-0.011) | <0.001 | 0.101 |
| | **rMSSD** | | | | | |
| | Two weeks before | 0.035(0.023,0.047) | <0.001 | 0.020(0.010,0.030) | <0.001 | 0.034 |
| | One week before | -0.028(-0.04,-0.016) | <0.001 | 0.001(-0.009,0.011) | 0.782 | <0.001 |
| | One week after | -0.044(-0.058,-0.03) | <0.001 | -0.007(-0.017,0.003) | 0.190 | <0.001 |
| | Two weeks after | -0.017(-0.029,-0.005) | 0.004 | -0.030(-0.038,-0.022) | <0.001 | 0.063 |
| | **HF** | | | | | |
| | Two weeks before | 0.042(0.03,0.054) | <0.001 | 0.024(0.014,0.034) | <0.001 | 0.015 |
| | One week before | -0.015(-0.027,-0.003) | 0.005 | 0.022(0.012,0.032) | <0.001 | <0.001 |
| | One week after | -0.021(-0.033,-0.009) | 0.002 | 0.001(-0.009,0.011) | 0.882 | 0.012 |
| | Two weeks after | -0.003(-0.015,0.009) | 0.566 | -0.023(-0.031,-0.015) | <0.001 | 0.006 |
| | **LF** | | | | | |
| | Two weeks before | 0.010(0.000,0.020) | 0.041 | 0.005(-0.003,0.013) | 0.189 | 0.425 |
| | One week before | -0.006(-0.016,0.004) | 0.271 | 0.015(0.007,0.023) | <0.001 | 0.001 |
| | One week after | -0.022(-0.034,-0.01) | <0.001 | 0.006(-0.002,0.014) | 0.150 | <0.001 |
| | Two weeks after | 0.004(-0.006,0.014) | 0.370 | -0.009(-0.017,-0.001) | 0.012 | 0.026 |
| | **LF/HF** | | | | | |
| | Two weeks before | -0.017(-0.027,-0.007) | 0.001 | 0.004(-0.004,0.012) | 0.376 | 0.001 |

| | | | | | | |
|---|---|---|---|---|---|---|
| | One week before | 0.011(0.001,0.021) | 0.036 | 0.002(-0.006,0.01) | 0.577 | 0.179 |
| | One week after | -0.006(-0.018,0.006) | 0.332 | 0.004(-0.004,0.012) | 0.423 | 0.209 |
| | Two weeks after | 0.015(0.005,0.025) | 0.002 | 0.008(0.000,0.016) | 0.042 | 0.206 |
| | **SNDD** | | | | | |
| | Two weeks before | 0.040(0.028,0.052) | <0.001 | 0.036(0.026,0.046) | <0.001 | 0.635 |
| | One week before | -0.016(-0.028,-0.003) | 0.012 | -0.004(-0.013,0.005) | 0.378 | 0.136 |
| | One week after | -0.050(-0.061,-0.039) | <0.001 | 0.019(0.010,0.028) | <0.001 | <0.001 |
| | Two weeks after | 0.001(-0.011,0.014) | 0.852 | -0.020(-0.027,-0.012) | <0.001 | 0.005 |
| | **rMSSD** | | | | | |
| | Two weeks before | 0.040(0.028,0.052) | <0.001 | 0.036(0.026,0.046) | <0.001 | 0.635 |
| | One week before | -0.016(-0.028,-0.003) | <0.001 | -0.004(-0.013,0.005) | 0.606 | 0.136 |
| | One week after | -0.050(-0.061,-0.039) | <0.001 | 0.019(0.010,0.028) | <0.001 | <0.001 |
| | Two weeks after | 0.001(-0.011,0.014) | 0.614 | -0.020(-0.027,-0.012) | <0.001 | 0.005 |
| | **HF** | | | | | |
| **Affective symptoms** | Two weeks before | 0.052(0.039,0.065) | <0.001 | 0.060(0.049,0.072) | <0.001 | 0.343 |
| | One week before | -0.016(-0.028,-0.003) | 0.015 | 0.004(-0.006,0.013) | 0.473 | 0.018 |
| | One week after | -0.051(-0.062,-0.039) | <0.001 | 0.029(0.019,0.039) | <0.001 | <0.001 |
| | Two weeks after | -0.005(-0.018,0.008) | 0.459 | -0.017(-0.026,-0.009) | <0.001 | 0.134 |
| | **LF** | | | | | |
| | Two weeks before | 0.031(0.02,0.043) | <0.001 | 0.016(0.007,0.026) | 0.001 | 0.051 |
| | One week before | -0.003(-0.015,0.009) | 0.604 | -0.004(-0.013,0.004) | 0.291 | 0.859 |
| | One week after | -0.035(-0.045,-0.024) | <0.001 | 0.013(0.005,0.022) | 0.001 | <0.001 |
| | Two weeks after | 0.005(-0.007,0.017) | 0.441 | -0.008(-0.015,-0.001) | 0.032 | 0.079 |
| | **LF/HF** | | | | | |
| | Two weeks before | -0.015(-0.027,-0.003) | 0.012 | -0.004(-0.014,0.005) | 0.359 | 0.163 |
| | One week before | 0.014(0.003,0.026) | 0.017 | -0.006(-0.014,0.003) | 0.170 | 0.006 |

| | | | | | | |
|---|---|---|---|---|---|---|
| | One week after | 0.004(-0.006,0.015） | 0.405 | 0.009(0.001,0.018） | 0.032 | 0.473 |
| | Two weeks after | -0.007(-0.019,0.004） | 0.224 | 0.003(-0.004,0.01） | 0.417 | 0.144 |

(Note: PMDs: premenstrual disorders, including PMS and PMDD. $B_1$: the coefficient of HRV-symptom associations among these with PMDs; $SE_1$: standardized error for $B_1$; $B_2$: the coefficient of HRV-symptom associations among these without PMDs; $SE_2$: standardized error for $B_2$; *P*-for-difference: *P* value for comparisons of $B_1$ and $B_2$. SDNN: standard deviation of RR intervals; rMSSD: Root mean square of successive RR interval differences; HF: absolute power in the HF band (0.15–0.4 Hz); LF: absolute power in the LF band (0.04–0.15 Hz); LF/HF: ratio of LF to HF. The models have been adjusted for age, age at menarche, BMI, drinking during the past 30 days, and drinking heavily during the past 30 days.)

**Table S3. Comparison of the associations between HRV and PMD symptoms among individuals with PMDD, PMS, and without PMDs across the menstrual cycle**

| Weeks | *PMDD* | | *PMS* | | *Non-PMDs* | | $P_1$-for-difference | $P_2$-for-difference | $P_3$-for-difference |
|---|---|---|---|---|---|---|---|---|---|
| | $B_1$ ( 95%CI) | *P* value | $B_2$ ( 95%CI) | *P* value | $B_3$ ( 95%CI) | *P* value | | | |
| SDNN | | | | | | | | | |
| Two weeks before | 0.049(0.033,0.065) | <0.001 | 0.034(0.016,0.052) | <0.001 | 0.028(0.02,0.036) | <0.001 | 0.213 | 0.019 | 0.542 |
| One week before | -0.047(-0.067,-0.027) | <0.001 | -0.012(-0.028,0.004) | 0.154 | 0.001(-0.007,0.009) | 0.720 | 0.006 | <0.001 | 0.146 |
| One week after | -0.05(-0.068,-0.032) | <0.001 | -0.048(-0.064,-0.032) | <0.001 | 0.007(-0.001,0.015) | 0.093 | 0.868 | <0.001 | <0.001 |
| Two weeks after | 0.000(-0.020,0.020) | 0.971 | -0.024(-0.042,-0.006) | 0.008 | -0.021(-0.029,-0.013) | <0.001 | 0.074 | 0.051 | 0.761 |
| rMSSD | | | | | | | | | |
| Two weeks before | 0.080(0.062,0.098) | <0.001 | 0.051(0.031,0.071) | <0.001 | 0.042(0.032,0.052) | <0.001 | 0.031 | <0.001 | 0.421 |
| One week before | -0.081(-0.103,-0.059) | <0.001 | -0.013(-0.031,0.005) | 0.128 | -0.001(-0.011,0.009) | 0.883 | <0.001 | <0.001 | 0.244 |
| One week after | -0.085(-0.105,-0.065) | <0.001 | -0.056(-0.074,-0.038) | <0.001 | 0.008(-0.002,0.018) | 0.106 | 0.031 | <0.001 | <0.001 |
| Two weeks after | 0.016(-0.006,0.038) | 0.144 | -0.057(-0.077,-0.037) | <0.001 | -0.032(-0.04,-0.024) | <0.001 | <0.001 | <0.001 | 0.02 |
| HF | | | | | | | | | |
| Two weeks before | 0.080(0.062,0.098) | <0.001 | 0.045(0.027,0.063) | <0.001 | 0.049(0.039,0.059) | <0.001 | 0.006 | 0.003 | 0.698 |
| One week before | -0.054(-0.076,-0.032) | <0.001 | 0.000(-0.018,0.018) | 0.988 | 0.014(0.006,0.022) | 0.002 | <0.001 | <0.001 | 0.155 |
| One week after | -0.036(-0.056,-0.016) | <0.001 | -0.047(-0.065,-0.029) | <0.001 | 0.017(0.007,0.027) | 0.001 | 0.414 | <0.001 | <0.001 |
| Two weeks after | 0.019(-0.001,0.039) | 0.069 | -0.044(-0.064,-0.024) | <0.001 | -0.021(-0.029,-0.013) | <0.001 | <0.001 | <0.001 | 0.033 |
| LF | | | | | | | | | |
| Two weeks before | 0.051(0.035,0.067) | <0.001 | 0.009(-0.009,0.027) | 0.334 | 0.012(0.004,0.02) | 0.003 | <0.001 | <0.001 | 0.761 |
| One week before | -0.019(-0.039,0.001) | 0.050 | 0.007(-0.009,0.023) | 0.388 | 0.006(-0.002,0.014) | 0.127 | 0.042 | 0.020 | 0.911 |
| One week after | -0.027(-0.045,-0.009) | 0.002 | -0.037(-0.053,-0.021) | <0.001 | 0.010(0.002,0.018) | 0.010 | 0.406 | <0.001 | <0.001 |
| Two weeks after | 0.010(-0.008,0.028) | 0.278 | -0.001(-0.019,0.017) | 0.935 | -0.009(-0.015,-0.003) | 0.007 | 0.387 | 0.045 | 0.399 |
| LF/HF | | | | | | | | | |

| | | | | | | | | | |
|---|---|---|---|---|---|---|---|---|---|
| Two weeks before | -0.012(-0.028,0.004) | 0.150 | -0.025(-0.043,-0.007) | 0.005 | 0.000(-0.008,0.008) | 0.940 | 0.280 | 0.180 | 0.011 |
| One week before | 0.020(0.000,0.040) | 0.043 | 0.024(0.008,0.04) | 0.004 | -0.001(-0.009,0.007) | 0.868 | 0.755 | 0.051 | 0.005 |
| One week after | 0.003(-0.015,0.021) | 0.714 | -0.003(-0.019,0.013) | 0.740 | 0.007(-0.001,0.015) | 0.111 | 0.618 | 0.685 | 0.264 |
| Two weeks after | -0.008(-0.026,0.01) | 0.364 | 0.027(0.009,0.045) | 0.003 | 0.006(0.000,0.012) | 0.081 | 0.006 | 0.140 | 0.027 |

(Note: PMDD: premenstrual dysphoric disorder; PMS: premenstrual syndrome; Non-PMDs: individuals without PMDD or PMS. $B_1$: the coefficient of HRV-symptom associations among these with PMDD; $SE_1$: standardized error for $B_1$; $B_2$: the coefficient of HRV-symptom associations among these with PMS; $SE_2$: standardized error for $B_2$; $B_3$: the coefficient of HRV-symptom associations among Non-PMDs; $SE_3$: standardized error for $B_3$. $P_1$-for-difference: comparisons between $B_1$ and $B_2$, $P_2$-for-difference: comparisons between $B_1$ and $B_3$, $P_3$-for-difference: comparisons between $B_2$ and $B_3$. SDNN: standard deviation of RR intervals; rMSSD: Root mean square of successive RR interval differences; HF: absolute power in the HF band (0.15–0.4 Hz); LF: absolute power in the LF band (0.04–0.15 Hz); LF/HF: ratio of LF to HF. According to the Bonferroni correction, the significance levels for $P_1$-for-difference, $P_2$-for-difference, and $P_3$-for-difference in the table have been adjusted to 0.05/3 = 0.017. The models have been adjusted for age, age at menarche, BMI, drinking during the past 30 days, and drinking heavily during the past 30 days.)

**Table S4. Comparison of the associations between HRV and PMD symptoms among individuals with PMDs and pure controls across the menstrual cycle**

| Weeks | PMDs | | Pure Control | | *P-for-difference* |
|---|---|---|---|---|---|
| | ***B*₁ ( 95%CI)** | ***P* value** | ***B*₂ ( 95%CI)** | ***P* value** | |
| SDNN | | | | | |
| Two weeks before | 0.035(0.023,0.046) | <0.001 | -0.015(-0.037,0.007) | 0.181 | <0.001 |
| One week before | -0.024(-0.035,-0.013) | <0.001 | -0.007(-0.028,0.015) | 0.544 | 0.175 |
| One week after | -0.05(-0.061,-0.039) | <0.001 | 0.005(-0.015,0.025) | 0.610 | <0.001 |
| Two weeks after | -0.004(-0.015,0.006) | 0.411 | 0.024(0.002,0.047) | 0.033 | 0.020 |
| rMSSD | | | | | |
| Two weeks before | 0.055(0.043,0.067) | <0.001 | -0.013(-0.037,0.011) | 0.286 | <0.001 |
| One week before | -0.036(-0.048,-0.025) | <0.001 | 0.02(-0.005,0.044) | 0.113 | <0.001 |
| One week after | -0.073(-0.084,-0.061) | <0.001 | 0.015(-0.008,0.039) | 0.207 | <0.001 |
| Two weeks after | -0.013(-0.025,-0.001) | 0.028 | 0.03(0.006,0.055) | 0.014 | 0.001 |
| HF | | | | | |
| Two weeks before | 0.054(0.042,0.066) | <0.001 | -0.026(-0.051,-0.001) | 0.045 | <0.001 |
| One week before | -0.021(-0.032,-0.009) | <0.001 | 0.012(-0.013,0.037) | 0.334 | 0.021 |
| One week after | -0.047(-0.059,-0.036) | <0.001 | 0.053(0.027,0.078) | <0.001 | <0.001 |
| Two weeks after | -0.005(-0.016,0.007) | 0.415 | 0.024(-0.002,0.049) | 0.067 | 0.043 |
| LF | | | | | |
| Two weeks before | 0.026(0.015,0.037) | <0.001 | -0.017(-0.037,0.004) | 0.106 | <0.001 |
| One week before | -0.006(-0.017,0.005) | 0.275 | -0.004(-0.024,0.016) | 0.703 | 0.858 |
| One week after | -0.035(-0.045,-0.024) | <0.001 | 0.02(0.002,0.039) | 0.032 | <0.001 |
| Two weeks after | 0.004(-0.006,0.014) | 0.481 | 0.009(-0.012,0.031) | 0.402 | 0.679 |
| LF/HF | | | | | |
| Two weeks before | -0.018(-0.029,-0.007) | 0.001 | -0.019(-0.041,0.002) | 0.083 | 0.936 |

| | | | | | |
|---|---|---|---|---|---|
| One week before | 0.016(0.005,0.027) | 0.003 | -0.045(-0.065,-0.025) | <0.001 | <0.001 |
| One week after | 0.002(-0.008,0.012) | 0.725 | 0.021(0.002,0.039) | 0.028 | 0.065 |
| Two weeks after | 0.003(-0.007,0.013) | 0.516 | -0.001(-0.022,0.02) | 0.929 | 0.741 |

(Note: PMDs: premenstrual disorders, including PMS and PMDD. SDNN: standard deviation of RR intervals; rMSSD: Root mean square of successive RR interval differences; LF: absolute power in the LF band (0.04–0.15 Hz); HF: absolute power in the HF band (0.15–0.4 Hz); LF/HF: ratio of LF to HF. The models have been adjusted for age, age at menarche, BMI, drinking during the past 30 days, and drinking heavily during the past 30 days.)

**Table S5. Comparisons of the associations between HRVs and PMD symptoms among individuals with PMDs and without PMDs across menstrual cycle in subpopulation with regular menstrual cycles**

| HRVs | *With PMDs* | | *Without PMDs* | | *P-for-difference* |
|---|---|---|---|---|---|
| | ***$B_1$* ( 95%CI)** | ***P value*** | ***B2* ( 95%CI)** | ***P value*** | |
| **SNDD** | | | | | |
| Two weeks before | 0.015(-0.001,0.031) | 0.077 | 0.044(0.028,0.06) | <0.001 | 0.031 |
| One week before | -0.014(-0.03,0.002) | 0.107 | 0.02(0.008,0.032) | <0.001 | 0.002 |
| One week after | -0.068(-0.084,-0.052) | <0.001 | 0.006(-0.006,0.018) | 0.355 | -0.052 |
| Two weeks after | 0.001(-0.013,0.015) | 0.884 | -0.038(-0.048,-0.028) | <0.001 | 0.015 |
| **rMSSD** | | | | | |
| Two weeks before | 0.007(-0.011,0.025) | 0.43 | 0.052(0.036,0.068) | <0.001 | 0.025 |
| One week before | -0.021(-0.039,-0.003) | 0.016 | 0.017(0.005,0.029) | 0.006 | -0.003 |
| One week after | -0.104(-0.12,-0.088) | <0.001 | -0.001(-0.015,0.013) | 0.836 | -0.088 |
| Two weeks after | -0.007(-0.023,0.009) | 0.369 | -0.048(-0.06,-0.036) | <0.001 | 0.009 |
| **HF** | | | | | |
| Two weeks before | 0.003(-0.015,0.021) | 0.706 | 0.059(0.041,0.077) | <0.001 | 0.021 |
| One week before | -0.015(-0.033,0.003) | 0.092 | 0.034(0.022,0.046) | <0.001 | 0.003 |
| One week after | <0.001 | <0.001 | 0.012(-0.002,0.026) | 0.098 | -0.063 |
| Two weeks after | -0.012(-0.028,0.004) | 0.104 | -0.039(-0.051,-0.027) | <0.001 | 0.004 |
| **LF** | | | | | |
| Two weeks before | 0.017(0.001,0.033) | 0.030 | 0.024(0.01,0.038) | 0.001 | 0.033 |
| One week before | 0.001(-0.015,0.017) | 0.882 | 0.021(0.011,0.031) | <0.001 | 0.017 |
| One week after | -0.041(-0.055,-0.027) | <0.001 | 0.013(0.001,0.025) | 0.024 | -0.027 |
| Two weeks after | 0.007(-0.007,0.021) | 0.355 | -0.016(-0.026,-0.006) | 0.001 | 0.021 |
| **LF/HF** | | | | | |
| Two weeks before | 0.015(-0.001,0.031) | 0.059 | 0.011(-0.003,0.025) | 0.124 | 0.031 |

| | | | | | |
|---|---|---|---|---|---|
| One week before | 0.029(0.011,0.047) | 0.001 | 0.003(-0.007,0.013) | 0.537 | 0.047 |
| One week after | -0.002(-0.016,0.012) | 0.770 | 0.016(0.004,0.028) | 0.007 | 0.012 |
| Two weeks after | -0.004(-0.018,0.01) | 0.548 | -0.005(-0.015,0.005) | 0.276 | 0.010 |

(**Note:** PMDs: premenstrual disorders, including PMS and PMDD. $B_1$: the coefficient of HRV-symptom associations among these with PMDs; $SE_1$: standardized error for $B_1$; $B_2$: the coefficient of HRV-symptom associations among these without PMDs; $SE_2$: standardized error for $B_2$; *P*-for-difference: *P* value for comparisons of $B_1$ and $B_2$. SDNN: standard deviation of RR intervals; rMSSD: Root mean square of successive RR interval differences; HF: absolute power in the HF band (0.15–0.4 Hz); LF: absolute power in the LF band (0.04–0.15 Hz); LF/HF: ratio of LF to HF. The models have been adjusted for age, age at menarche, BMI, drinking during the past 30 days, and drinking heavily during the past 30 days.)
